# Clinical validation of RNA sequencing for Mendelian disorder diagnostics

**DOI:** 10.1101/2024.08.15.24312057

**Authors:** Sen Zhao, Kristina Macakova, Jefferson C. Sinson, Hongzheng Dai, Jill Rosenfeld, Gladys E. Zapata, Shenglan Li, Patricia Ward, Christiana Wang, Chunjing Qu, Becky Maywald, Undiagnosed Diseases Network, Brendan Lee, Christine Eng, Pengfei Liu

## Abstract

Despite rapid advancements in clinical sequencing, over half of diagnostic evaluations still lack definitive results. RNA-seq has shown promise in research settings for bridging this gap by providing essential functional data for accurate interpretation of diagnostic sequencing results. However, despite advanced research pipelines, clinical translation of diagnostic RNA-seq has not yet been realized. We have developed and validated a clinical diagnostic RNA-seq test in a CLIA laboratory for individuals with suspected genetic disorders who have existing or concurrent comprehensive DNA diagnostic testing. This diagnostic RNA-seq test processes patient RNA samples from fibroblasts or blood and derives clinical interpretations based on the analytical detection of outliers in gene expressions and splicing patterns. The clinical validation involves 150 samples, including benchmark, negative, and positive samples. We developed provisional expression and splicing benchmarks using short-read and long-read RNA-seq data from the HG002 lymphoblastoid sample produced by the Genome in a Bottle Consortium. Our validation data achieved analytical sensitivity and specificity higher than 99% against the benchmarks. For clinical validation, we first established reference ranges for each gene and junction based on expression distributions from our control data. We then evaluated the clinical performance of our outlier-based pipeline using positive samples with previously identified diagnostic findings from the Undiagnosed Diseases Network project. Our pipeline identified 19 of 20 positive findings in both fibroblast and blood samples and highlighted the limitations of the test. Our study provides a paradigm and necessary resources for independent laboratories to validate a clinical RNA-seq test.

## Introduction

The rapid advancements of sequencing technologies, such as clinical exome sequencing (ES) and whole genome sequencing (GS), have revolutionized the diagnosis of Mendelian disorders in the past decade ^1,2^. Meanwhile, the implementation of genome-wide NGS has led to the identification of numerous variants with unknown impacts on RNA and protein, which brings challenges to clinical interpretation ^3^. Recently, transcriptome RNA sequencing (RNA-seq) has emerged as a powerful adjunct to ES and GS in Mendelian disorder diagnostics ^4^. Owing to its ability to detect abnormal expression and splicing patterns, transcriptome sequencing can improve molecular diagnostic rates by 7.5% to 36% compared with DNA testing alone ^5–8^. Despite the development of advanced research pipelines by various groups, the clinical translation of diagnostic RNA-seq has not yet been realized. The field lacks the comprehensive implementation knowledge required for such a transition.

Several guidelines have been established for the clinical validation of NGS-based DNA sequencing ^9–13^. While the principles and experiences from DNA NGS tests can guide RNA-seq validation, key differences add to the challenges of validating RNA-seq tests. Despite originating from the same type of sequencing read raw data, DNA-based NGS and RNA-seq diverge in their analytical endpoints. DNA sequencing typically focuses on detecting SNVs/INDELs or copy number variants (CNV), whereas RNA-seq aims to measure gene expression levels and splicing junction status. Therefore, efforts are needed to adapt the DNA-based validation framework to RNA-seq analytical endpoints. Benchmark data should be generated, preferably using publicly accessible resources, such as those from the Genome in a Bottle Consortium (GIAB) consortium from the National Institute of Standards and Technology (www.nist.gov/programs-projects/genome-bottle), to facilitate seamless adoption for diagnostic labs in implementing this validation.

Both DNA- and RNA-based diagnostic tests are designed to identify rare genetic findings that explain the rare disease phenotype. The population distribution of DNA variants typically follows a zero-inflated bimodal distribution, where a significant number of observations occur at zero (i.e., reference genotype). This characteristic facilitates effective cross-platform data comparison, enabling the use of external large control databases, such as the gnomAD ^14^, thus reducing the burden on clinical labs to produce control data. In contrast, both gene expression and junction splicing data from RNA-seq exhibit a wide distribution in the “normal” population, increasing the challenge of distinguishing diagnostic outliers from background noise. Therefore, it is crucial for RNA-seq tests, especially in the clinical context, to generate control data using the same experimental and bioinformatics pipeline. Additionally, there is a strong need to establish reference ranges for all targets in the RNA-seq test, a component that is usually less extensively considered when validating DNA-based NGS tests.

Tissue-specific expression of genes and transcripts presents another challenge for the design and clinical validation of the RNA-seq tests. According to GTEx, 37.4% and 48.3% of all coding genes are low-expressing (TPM < 1) in blood and fibroblast tissues, respectively, which are the most frequently used clinically accessible tissues ^15,16^. This expression heterogeneity suggests that most diagnostic RNA-seq validations should be designed in a tissue-dependent manner to address test performance and clinical limitations.

Here, we report the clinical validation processes of a diagnostic RNA-seq test for the diagnosis of Mendelian disorders. We included publicly available benchmark samples, clinically positive samples, and negative control samples. We conducted optimization and familiarization (O&F) and set up passing criteria for quality control. We set up a provisional RNA standard reference using data from the GIAB consortium and evaluated the analytical performance of our test. Additionally, we established transcriptome-wide reference ranges for all reportable targets. Finally, we assessed the clinical performance using positive samples with previously identified diagnostic findings from the Undiagnosed Diseases Network (UDN) project.

## Material and methods

### Ethics approval

This study has been approved by the Institutional Review Board (IRB) at Baylor College of Medicine (H-42680). The study subjects were originally recruited through informed consent approved the IRB at the National Human Genome Research Institute (15HG0130) or BCM (H-34433 and H-44172), and then de-identified for the current study.

### Collection of Validation sample

A total of 130 samples were collected from 110 individuals, including 73 fibroblast samples, 55 blood samples, and 2 lymphoblastoid/lymphocyte samples (**Table 1**). None of the individuals are related to each other.

Positive samples from UDN were selected under the following criteria:

1. A molecular diagnosis of a Mendelian disorder supported by DNA variant(s).
2. DNA variant(s) predicted to result in RNA-level changes (altered expression or alternative splicing).
3. Prior RNA-seq studies identified the predicted RNA-level changes.
4. At least two independent experimental data sources support the result.

Negative samples from the UDN were selected based on the following criteria:

1. No known molecular diagnosis of a Mendelian disorder.
2. No recorded kinship to other negative or positive samples in the cohort.

### Sample processing and RNA extraction

Fibroblast samples were cultured in high-glucose DMEM medium, supplemented with 10% fetal bovine serum (FBS), 1% non-essential amino acid (NEAA), and 1% penicillin-streptomycin (P-S). Blood samples were stored in PAXgene tubes at -80 before RNA extraction. RNA was extracted from around 1×10^7^ cells using the RNeasy mini kit (Qiagen) following the manufacturer’s instructions with the inclusion of an on-column genomic DNA removal step. The integrity and quality of the RNA were assessed using the Qubit 4 Fluorometer and the Qubit RNA HS Assay Kit (ThermoFisher).

### Library preparation and NGS

RNA from fibroblast and lymphoblastoid/lymphocyte samples was processed using the Illumina Stranded mRNA prep kit. RNA from whole-blood samples was processed using the Illumina Stranded Total RNA Prep with Ribo-Zero Plus kit to remove human globin RNA and rRNA. Sequencing was conducted on the Illumina NovaSeqX platform, which produces pared-end short read data at 150bp. Each validation sample was sequenced on average to a target depth of 150 million reads.

### Raw data processing and expression quantification of RNA-seq

The raw data processing pipeline was adapted from the Genotype-Tissue Expression (GTEx) version 10 pipeline (https://github.com/broadinstitute/gtex-pipeline/blob/master/rnaseq/README.md). Sequencing FASTQ data were aligned to the reference genome GRCh38 with STAR v2.7.8a_sentieon and SAMtools 1.15.1/HTSlibv1.10.2. Picard v2.23.3 was used to mark duplicates. Gene expressions were quantified using RNA-SeQC v2.4.2 ^17^. Isoform-level quantification was performed with RSEM v1.3.3 ^18^. Transcripts were annotated with GENCODE v39. FastQC v0.11.9 provided quality control measurements. For identity verification, SNP/indels were called from the RNA sequencing data using the haplotyper from Sentieon DNAseq. The resultant variants were compared to those obtained from DNA sequencing data of the same individual to ensure identity matching.

### Reproducibility test

Using a 3-1-1 validation framework, a reproducibility test was performed on HG002, K562, and BG1477. We conducted an intra-run with triplicate preparations of the same sample, followed by two inter-runs of the same sample, resulting in a total of five tests across three different batches prepared on three different days. The intra-run and inter-run batches were prepared by two different technicians using different Bio-Rad thermocyclers, pipettes, vorterxers, centrifuges and reagents. This comprehensive approach enabled us to measure resulting variations, affirming the assay’s consistent and reliable performance.

Gene expression reproducibility was calculated by performing pair-wise Pearson correlation of gene read counts. Only genes with adequate expression (TPM > 5) were included. Splice junction detection reproducibility was calculated by pair-wise Pearson correlation of junction reads. Only canonical junctions in genes with adequate expression (TPM > 5) were included.

### RNA-seq Benchmark data

GM24385 is a human lymphoblastoid cell line derived from a female donor HG002, widely used for benchmarking and validating DNA-based genomic analyses due to its easy accessibility and extensive characterization ^19–23^. However, a ‘gold standard’ benchmark has not yet been established at the transcriptome level. Therefore, we sought to create a provisional benchmark by aggregating sequencing data generated independently from renowned groups from the GIAB team. The analysis incorporated data from different institutions, produced in multiple runs using both short- and long-read sequencing technologies. We obtained sequencing data that are released by the GIAB team^24^.

For short-read data, four sequence datasets from two groups were used:

1. UNC Data: A triplicate set of Illumina short-read RNA-seq data from The University of North Carolina (UNC). This data was generated on three HG002 cell lines as part of the NIST-GIAB RNAseq pilot sequencing project (https://ftp-trace.ncbi.nlm.nih.gov/ReferenceSamples/giab/data_RNAseq/AshkenazimTrio/HG002_NA24385_son/UNC_Illumina/).
2. Google Data: Another set of Illumina short-read RNA-seq data provided by Google, with sequencing contracted out to Novogene as part of the NIST-GIAB RNA-seq pilot sequencing project (https://ftp-trace.ncbi.nlm.nih.gov/ReferenceSamples/giab/data_RNAseq/AshkenazimTrio/HG002_NA24385_son/Google_Illumina/).

Short-read benchmark data were processed using the same pipeline mentioned above. We first averaged the TPM of protein-coding genes within the three UNC datasets and then further averaged the result with the Google sample to create a standard mean TPM matrix. Genes with mean TPM ≥ 5 were selected as the positive expression set, while genes with mean TPM = 0 as the negative expression set. This expression benchmark included 8991 positive genes and 1296 negative genes. To validate our in-house data, a gene was defined as detected if the read count was ≥ 50, and undetected if the read count was < 50. Sensitivity (positive rate of genes in the positive expression set) and specificity (negative rate of genes in the negative expression set) were used to evaluate the performance of gene expression quantification.

For long-read data, two sequence datasets from two groups were used:

1. PacBio Data: PacBio long-read RNA-seq data produced from the Sequel® Revio system by PacBio, using the Kinnex full-length RNA protocol (https://downloads.pacbcloud.com/public/dataset/Kinnex-full-length-RNA/DATA-Revio-HG002-1/).
2. Baylor Data: PacBio long-read RNA-seq data produced by Baylor College of Medicine, with sequencing contracted out to Novogene as part of the NIST-GIAB RNA-seq pilot sequencing project (https://ftp-trace.ncbi.nlm.nih.gov/ReferenceSamples/giab/data_RNAseq/AshkenazimTrio/HG002_NA24385_son/). Iso-Seq SMRT® libraries were constructed and sequenced using the PacBio Sequel Systems.

The Iso-Seq workflow (v4.0.0, https://isoseq.how/) was used for data processing. Reads were first mapped to hg38 using pbmm2 v1.13.1. Mapped reads were then collapsed into unique isoforms using isoseq v4.0.0, and classified and filtered using pigeon v1.1.0.

A provisional benchmark for splicing junctions was constructed by intersecting junctions passing quality control from each source. The consensus list was filtered for canonical junctions represented in GENCODE v39. To avoid detection noises from genes with insufficient expression in the LCL sample, only junctions mapped to genes with an adequate expression (TPM ≥ 5 according to GIAB short-read data) were used as the positive splicing set. Canonical junctions in genes from the negative expression set were used as the negative splicing set. This splicing benchmark included 38110 positive junctions and 4195 negative junctions. To validate our in-house data, a junction was defined as detected if the junction read count was ≥ 5, and undetected if the junction read count was < 5. Sensitivity (positive rate of junctions in the positive set) and specificity (negative rate of junctions in the negative set) were used to evaluate the performance of splicing junction detection.

### Reference ranges, outlier analysis, and clinical performance evaluation

We established a reference panel consisting 73 fibroblast samples and 55 blood samples from the validation cohort. Outlier analyses were performed independently for fibroblast samples and blood samples.

The expression outlier pipeline was adapted from Outrider v1.17.2 ^25^. A negative binomial distribution was modeled based on the raw counts of the gene across all samples using the R package MASS v7.3-60.0.1. Expression outliers were identified by comparing the expression value of each gene in each sample against this distribution. The range of expression folds corresponding to a P-value < 0.05 was defined as the reference range of a gene.

The splicing outlier pipeline was adapted from FRASER v1.99.1 ^26^. We modeled the percent spliced-in (PSI) value of each junction across the reference panel into a beta-binomial distribution using the R package VGAM v1.1-9. Splicing outliers were identified by comparing the PSI of each junction in each sample against this distribution. The range of PSI corresponding to a P-value < 0.05 was defined as the reference range of a junction.

For clinical performance, we evaluated the concordance of the outlier results with the known expression or splicing abnormalities in the positive samples. For expression abnormalities, the direction of change (up or down-regulation) and P-value from Outrider were used to determine whether the expected expression abnormality was detected. For splicing abnormalities, a splice-altering variant may result in changes in splicing conditions at multiple loci within a gene. For example, a DNA variant causing exon skipping leads to alteration of two splice conditions: a reduction of PSI for the intron proceeding and the intron following the skipped exon. In our clinical validation, positive detection of splicing abnormality was defined as having at least one of the expected PSI changes flagged by the pipeline. Sensitivity analysis was performed by using different P-value cutoffs to filter the outlier results. At each P-value cutoff, the number of outliers and the number of positive cases were calculated.

## Results

### Design of a clinical transcriptome sequencing test and scope of the clinical validation

We developed a clinical RNA-seq test for the diagnosis of Mendelian disorders. This test is recommended for individuals with suspected genetic disorders who are undergoing or have completed a comprehensive DNA analysis, such as large panel testing, ES, or GS. Acceptable sample types include blood or cultured fibroblast samples, or total RNA extracted from these two sample types.

The test workflow is depicted in **Figure 1**. For fibroblast samples, a poly-A enrichment library is prepared, while for blood samples, a ribo depletion library is used. RNA-seq is performed on the Illumina platform, targeting a depth of 150 million reads. The raw data is processed using an in-house analytic pipeline adapted from the GTEx version 10.

**Figure 1.**
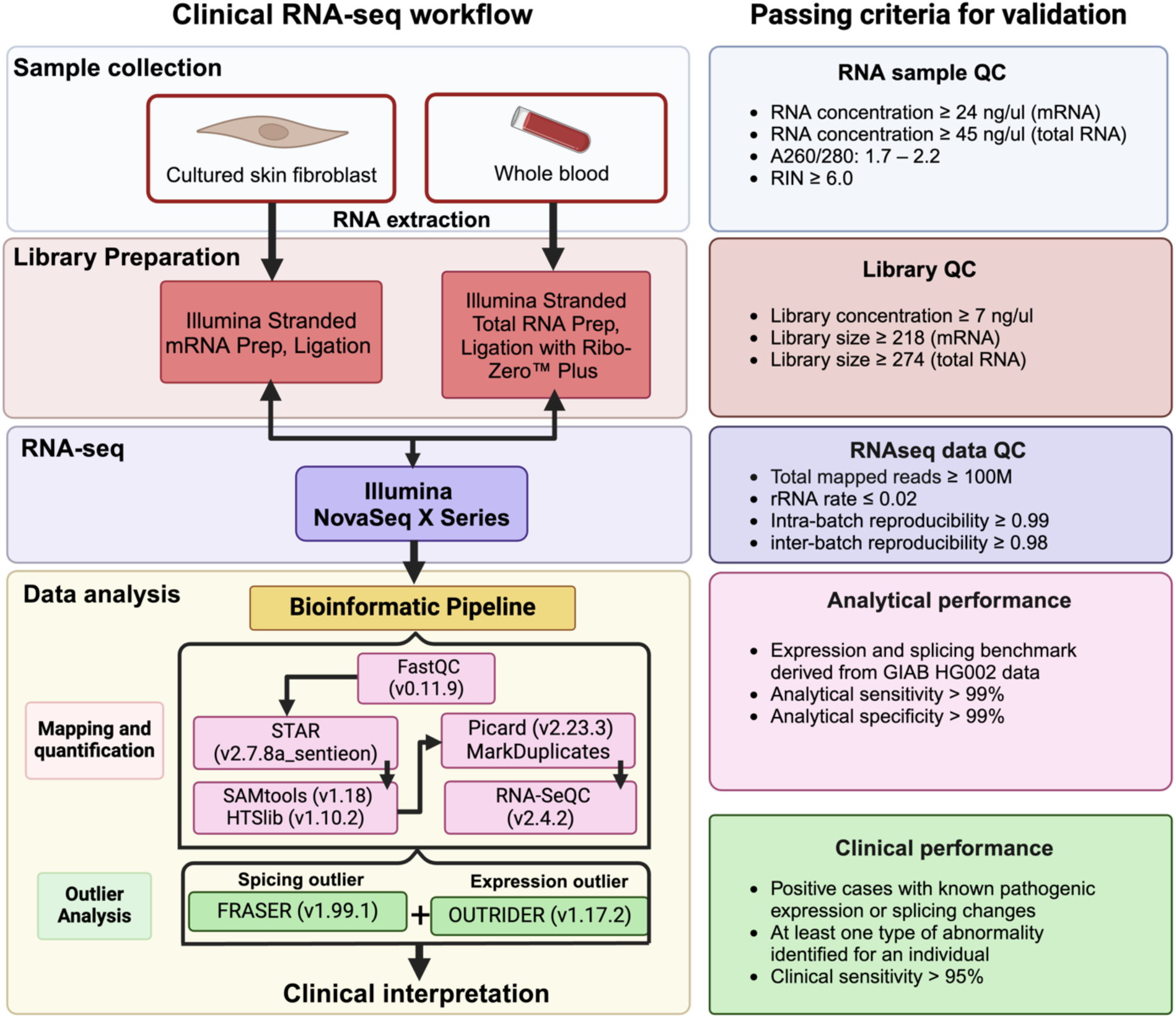
The workflow of clinical RNA-seq. The procedures of clinical RNA-seq is shown along with the passing criteria set for the validation of the test.

An outlier-based pipeline is used to identify abnormal expression and splicing events, using fixed reference data for blood (n=55) and fibroblast (n=73) samples. The abnormal expression and splicing events are interpreted in combination with DNA variants and patient phenotypes by American Board of Medical Genetics and Genomics (ABMGG)-certified professionals (**Figure S1**). The interpretation results and clinical recommendations are reported within a turnaround time of eight weeks.

The validation workflow incorporates an essential preliminary step of optimization and familiarization (O&F), followed by the formal validation. The formal validation phase involves selecting validation samples, formulating quality control (QC) metrics, testing reproducibility, evaluating analytical performance, establishing reference ranges, and assessing clinical performance.

### Cohort sample selection

A total of 130 samples from 110 individuals were included in the validation workflow, including 73 fibroblast samples, 55 blood samples, and 2 lymphoblastoid/lymphocyte samples (**Table 1)**. None of the individuals within each sample cohort are related to each other.

All 73 fibroblast samples were obtained from the Undiagnosed Diseases Network (UDN), comprising 20 positive samples with a molecular diagnosis and 43 ‘negative’ samples from apparently healthy siblings or parents of unrelated participant families. Ten of the negative samples were utilized for optimization and familiarization (O&F). Among the 55 blood samples, 54 were sourced from the UDN and one from a healthy volunteer (BG1477). The UDN blood samples included 20 positive samples and 34 ‘negative’ samples from apparently healthy siblings or parents of unrelated participant families, with 10 negative samples designated for O&F. The blood sample from BG1477 was used for reproducibility testing.

The two lymphoblastoid/lymphocyte samples, GM24385 and K562, were procured from the Coriell Institute. GM24385 (HG002) is a standard reference sample from the GIAB project and was used for both reproducibility testing and analytical validation. K562, derived from the bone marrow of a patient with chronic myeloid leukemia, was used for reproducibility testing.

### Quality controls

Prior to the formal validation, an O&F procedure was conducted. This involved calibrating equipment, testing reagents, establishing protocols, and formulating QC standards. Using O&F samples, we assessed the quality spectrum of RNA and NGS libraries (**Figure 2A**), and determined QC standards for the clinical test (**Figure 1**). The RNA sequencing yield was empirically targeted at 150 million reads (**Figure 1)**. Following raw data analysis, QC standards for the RNA-seq results were implemented based on statistics from the O&F data (**Figure 1**, **Figure 2B**). Identity matching was conducted for every sample as a QC procedure to prevent sample swap by comparing the concordance of SNPs identified from RNA-seq data with variant calls from existing DNA sequencing data.

**Figure 2.**
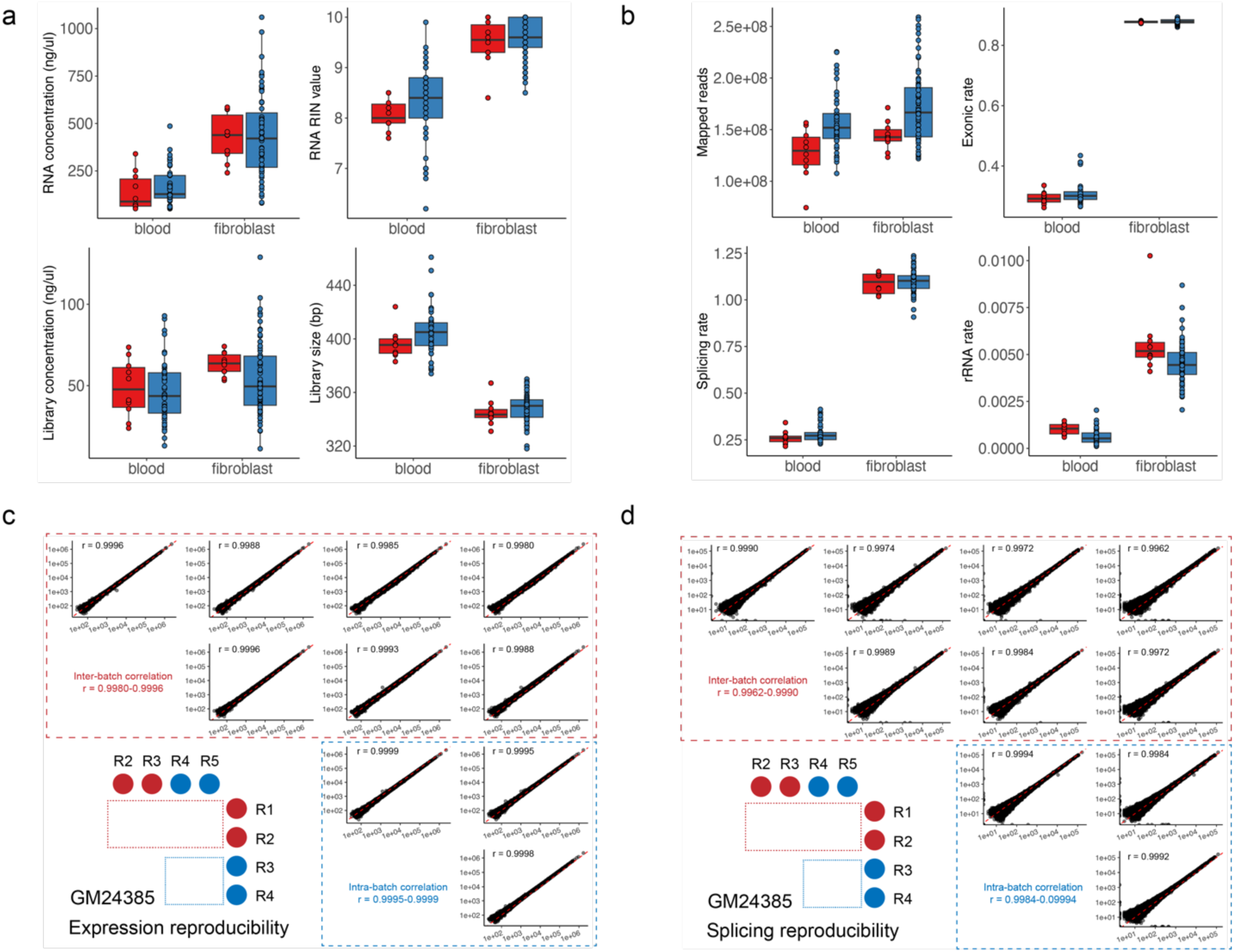
Quality control and reproducibility tests for pre-analytical sample, post-analytical library, and RNA-seq data. (A) RNA and library quality control metrics for obtained from Optimization and Familiarization (O&F, red) and other validation (blue) runs. (B) RNA-seq data quality control metrics of O&F (red) and other validation runs (blue). (C) Reproducibility tests for gene expression and splicing (D) metrics for GM24385. Intra-batch comparisons are boxed in blue dotted lines, whereas inter-batch comparisons are boxed in red.

### Reproducibility test

The reproducibility analysis was conducted to evaluate the consistency of results across repeat runs. The setup included five replicates: one triplicate in a batch and two individual runs in two other batches, enabling intra- and inter-batch reproducibility evaluation. We decided to calculate reproducibility using two fundamental measurements from RNA-seq results: gene expression levels and splicing status at exon-intron junction sites. Three distinct samples were included in this analysis: GM24385 (a lymphoblastoid cell line from HG002), K562 (a lymphoblast cell line), and BG1477 (a blood sample).

The expression-level correlation between each pair of replicates was calculated using the read count of coding genes. The splicing-level correlation between each pair of replicates was calculated using the read counts at GENCODE canonical junctions in genes. Genes with low expression levels (TPM < 5) were excluded from both correlation analyses. For all three samples, high reproducibility was achieved, with intra-batch correlations (Pearson coefficient) above 0.99 and inter-batch correlations (Pearson coefficient) above 0.98 (**Figure 2C, Figure S2, Figure S3**).

### Validation of analytical performance

The validation of a diagnostic test’s analytical performance typically involves assessing the detection of fundamental analytical elements. As an analogy, DNA-based NGS often measures the accuracy of base calling using gold standard samples. In the context of RNA-seq, we posit that the detection of gene expression levels and splicing status at exon-intron junctions should be extensively characterized.

We used GM24385, the lymphoblastoid cell line from the GIAB reference individual HG002 for our evaluation. We established expression and splice junction benchmarks for GM24385 using publicly available RNA-seq data. The expression benchmark was constructed using four short-read RNA-seq datasets from two independent laboratories. We calculated the mean TPM for each protein-coding gene between two laboratories and selected 8552 positive genes (TPM ≥ 5) and 1320 negative genes (TPM=0) (**Data S1**). The splicing benchmark was constructed using two long-read RNA-seq datasets from two independent laboratories. Positive junctions were defined as canonical junctions in the aforementioned positive gene set detected in both long-read datasets. Negative junctions were defined as canonical junctions in the negative gene set that were not detected in either long-read dataset. Based on these criteria, we identified 38110 positive junctions and 4195 negative junctions (**Data S2**). The positive and negative genes/junctions were then used as a reference to calculate the sensitivity and specificity of our test.

At the expression level, the five GM24385 replicates yielded a sensitivity ranging from 99.9% to 99.93% and a specificity of 100%. Most false negative (FN) genes exhibited apparent discrepancies in expression levels between the two benchmark resources, suggesting an impact of laboratory context on RNA-seq results (**Data S3**). This finding underscores the importance of including data from multiple sources and warrants further investigation. At the splicing level, the sensitivity ranged from 99.64% to 99.78%, and the specificity ranged from 99.79% to 99.9%. Notably, none of the FN junctions overlapped with the expression-level FN genes, suggesting that FN junctions were driven by alternative splicing patterns rather than inadequate gene expression. This observation was confirmed by manual inspection (**Data S4**, **Figure S4**). In contrast, false positive (FP) junctions were mostly attributed to mapping issues (**Data S4**, **Figure S5**). Overall, the analytical performance of our test exceeded the preset cutoff of 99% for both sensitivity and specificity at the expression and junction levels.

### Establishing transcriptome-wide reference ranges

The detection of outliers relies on the definition of a reference range, which is the range of values for given targets expected in a healthy population. For this transcriptome test, we first aimed to define the scope of genes sufficiently interrogated by the assay for outlier analysis and then establish reference ranges for every gene within this scope. Data from a reference panel of 73 fibroblast samples and 55 blood samples from the validation cohort were used to establish the reportable targets and the reference ranges.

Among all 19216 coding genes, 11962 (62.2%) from blood and 12614 (65.6%) from the fibroblast are deemed reportable (**Data S6** and **S7**), with a minimum of 50 sequencing reads from the average of all samples. When breaking down the gene list into disease-specific subsets, blood RNA-seq demonstrated gene coverage ranging from 56.8% to 85.4%, whereas fibroblast RNA-seq ranged from 67.3% to 86.7% (**Figure 3A)**. Consistent with previous reports ^6,27^, fibroblast demonstrated higher gene coverage compared to blood for all disease-specific panels except for immunodeficiency.

**Figure 3.**
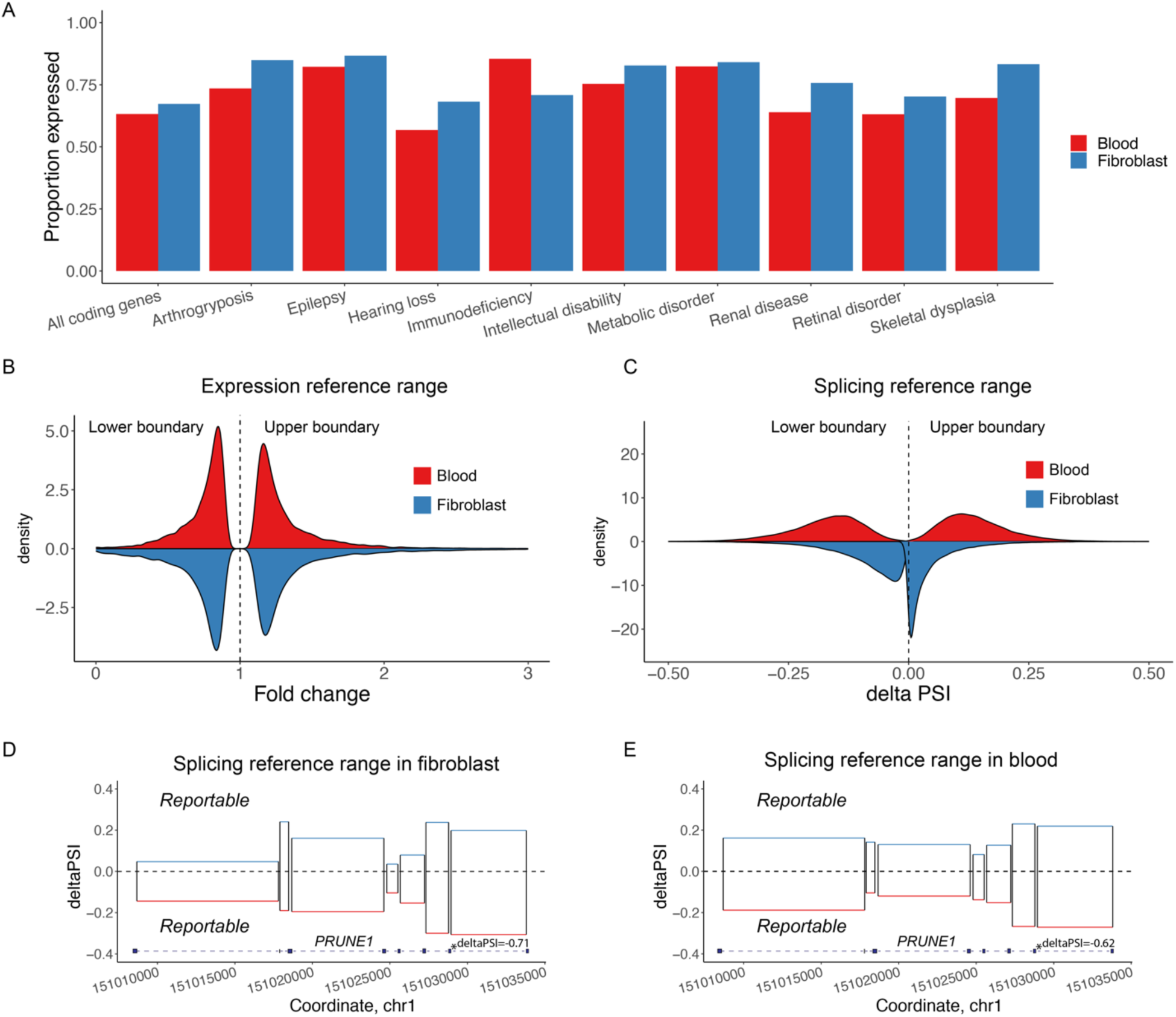
Characterization of transcriptome-wide expression profiles and reference ranges. (A) The gene expression profiles were based on the mean expression value of our fibroblast and blood reference cohorts. The proportions of genes with read counts ≥ 50 among all coding genes (n=19216) and are shown across various disease-specific gene panels. The distribution of transcriptome-wide reference ranges for gene expressions are plotted (B) and those for splicing are plotted in (C). The fold change/delta PSI values for the lower and the higher boundaries for each gene/junction are depicted on the graph. The range in between the lower and higher boundaries represent the reference range, while the ranges outside show the extent of abnormal changes that can be detected. Genes and junctions with low expressions are excluded from this analysis, resulting in the following number of targets in each assay: expression in blood n = 11962; expression in fibroblast n = 12614; splicing in Blood n = 105114; splicing in fibroblast n= 119403. The splicing reference ranges of *PRUNE1* is illustrated as an example based on our fibroblast (D) and blood (E) tests.

To establish expression reference ranges, we modeled the read count of each gene across the reference panel into a negative binomial distribution. The upper and lower boundaries of the range of expression fold changes corresponding to a P-value < 0.05 were defined as the reference range of a gene (**Data S6 and S7**, **Figure 3B**). The distribution of expression fold changes for all reportable genes showed mode values of 0.83 to 1.17 for blood and 0.85 to 1.16 for fibroblasts. This analysis suggests that the primary difference in gene expression levels between blood and fibroblast lies in the number of reportable genes, rather than the distribution of reference ranges among those reportable genes.

To establish splicing reference ranges, we modeled the percent spliced in (PSI) of each junction across the reference data using a beta-binomial distribution. The range of deltaPSI corresponding to a P-value < 0.05 was defined as the reference range of a junction (**Data S8 and S9, Figure 3C**). As an example, the splicing reference ranges for all splice junctions in *PRUNE1* are shown in **Figure 3D/E**.

After excluding canonical introns with fewer than three junctions in more than 20% of samples, 119403 and 105114 canonical introns remain for splicing reference range analysis in fibroblast and blood, respectively. The splicing reference range pattern differs dramatically between the two sample types. The distribution of fibroblast junction PSI shows a narrow reference range (equivalent to a wide outlier detection range) with mode values at -0.003 to 0.0002, compared to the mode reference range of -0.13 to 0.1 for blood. This difference is at least partially attributed to the distinct library preparation methods used. The polyA enrichment protocol for fibroblast boosts the proportion of canonical splicing in mature mRNA, leading to lower variation in PSI. In contrast, the ribo-depletion protocol for blood preserves unspliced pre-mRNA, resulting in higher PSI variation.

### Validation of clinical performance

Clinical performance is defined as the detection rate of clinical deliverables in an evaluation. For our clinical transcriptome analysis, we considered outliers for gene expression level or splice junction PSI corresponding to diagnostic DNA variants as the two key components that constitute the clinical deliverable. We calculated standalone detection rates for each of the two outlier types. Additionally, we determined patient-level detection rates by requiring the detection of only one outlier type when both expression and splicing outliers are expected in a single patient. This approach is justified because, when two outlier types are predicted from one event, it typically involves cryptic splicing that leads to nonsense-mediated decay, resulting in reduced expression levels. The intensity of residual cryptic splicing and the degree of gene expression reduction tend to counterbalance each other.

Positive clinical samples from the UDN, comprising 20 blood samples and 20 fibroblast samples from 23 individuals, were selected for the evaluation. These patients carry diagnostic variants encompassing a wide range of variant types, including putative loss-of-function variants, splicing variants, and deletions. The resultant RNA changes span a full spectrum including aberrant expression at the gene level, and aberrant splicing in the form of cryptic splice sites, exon skipping, cryptic exons, and fusion genes. To ensure the authenticity of these positive findings, all samples required at least two independent sets of experimental data affirming the diagnostic finding. The DNA variants, expected RNA findings, and descriptions of the supportive evidence are listed in **Data S5**. In some individuals, both expression and splicing abnormalities were expected, while in others, only one type of abnormality was anticipated. For our clinical validation, at least one type of abnormality was required to be detected in at least 95% of patients, meaning no more than one false negative was allowed for each sample type (**Figure 1)**.

For fibroblast samples, outlier analyses identified an average of 1060 expression outliers and 3910 splicing outliers per sample with P values < 0.05. In blood samples, more outliers were detected, with 1182 expression outliers and 6920 splicing outliers. We identified 12 out of 13 expected expression abnormalities and 16 out of 17 splicing abnormalities in fibroblast samples (**Data S5, Data S10, Figure 4A).** For combined sample-level findings, 19 out of 20 fibroblast samples showed at least one outlier abnormality. In blood samples, we identified 10 of 12 expected expression abnormalities and 15 of 17 splicing abnormalities, leading to a combined detection rate of 19 out of 20 samples (**Data S5, Data S11, Figure 4B)**. Both sample types achieved a sample-level detection rate of 95% (19 out of 20), meeting the preset requirement (**Figure 1**). The missing molecular diagnoses were attributed to two main factors: high background noise obscuring the low-impact positive findings (low expression of *AP4M1* in blood, **Figure S6;** low reduction of *PPP3CA* expression in fibroblast, **Data S10**), and the apparently variable efficiencies of nonsense-mediated mRNA decay (low NMD for *AP4M1* expression in blood, **Figure S6**).

**Figure 4.**
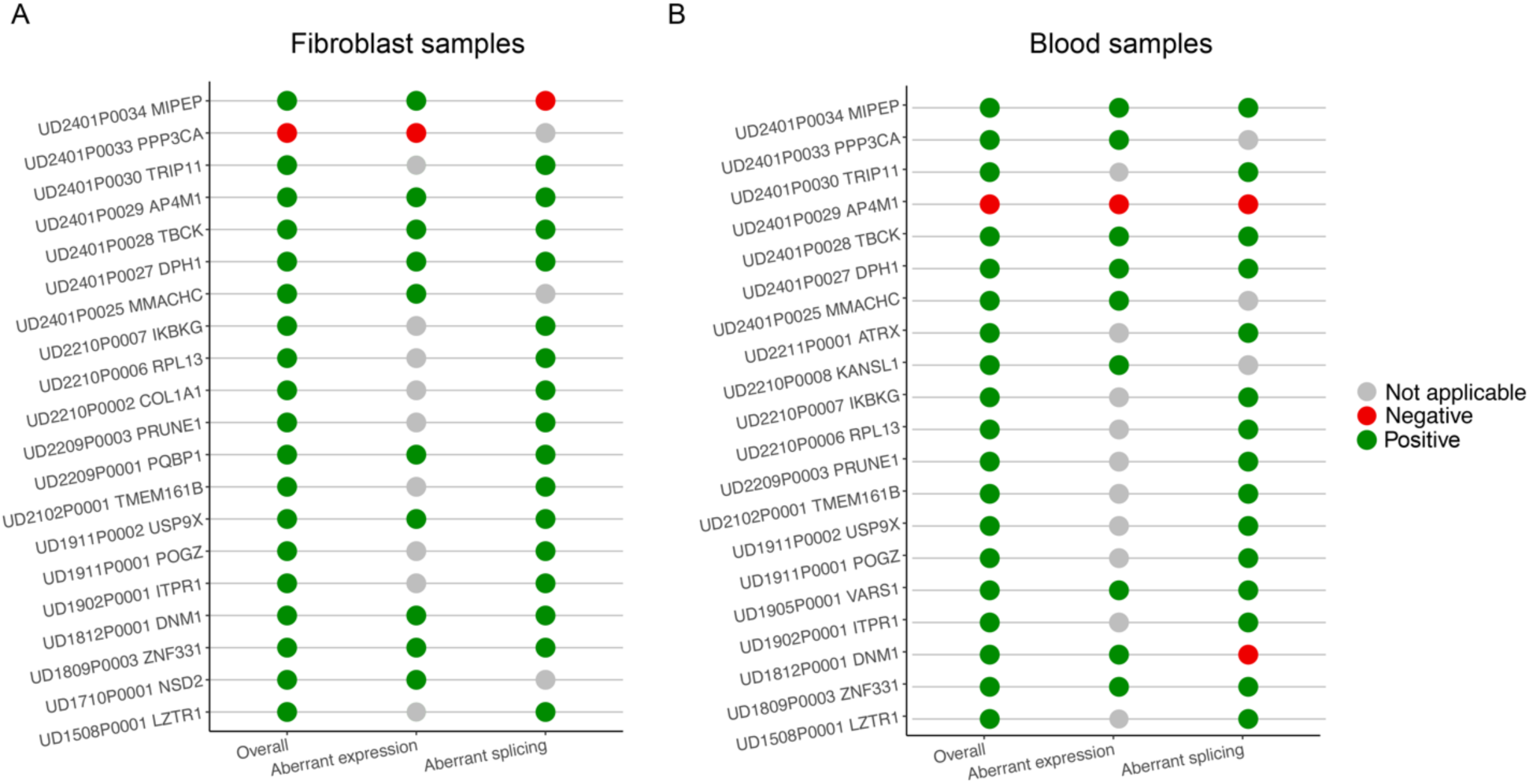
Clinical performance validation. Clinical performance in fibroblast samples (A) and blood samples (B). We evaluated whether each expression/splicing change was detected by our clinical pipeline. ‘Not applicable’ indicates that the expression/splicing change in this gene is not expected.

We then investigated how adjusting P-value cutoffs affects the number of outliers to review (reflecting positive predictive value) and the number of positive molecular diagnoses identified (reflecting sensitivity). Our findings indicate that fibroblast data are more robust against stringent P-value cutoffs. In fibroblast samples, most positive findings remain detectable even when cutoff stringencies are raised to retain approximately 100 reviewable expression or splicing outliers (**Figure S7**). In contrast, applying the same filtering criteria to blood samples results in the loss of more than half of the positive findings (**Figure S7**). It is crucial for clinical laboratories to balance clinical sensitivity with the interpretation workload and make informed decisions regarding filtration cutoffs for reviewable data.

## Discussion

Here, we report the development and clinical validation of an RNA-seq test for the diagnosis of Mendelian disorders. We demonstrate that a clinical workflow can be established to achieve high clinical sensitivity and specificity. Additionally, we also provide essential resources and considerations for conducting clinical validation, along with metrics necessary to define test limitations in the validation and to monitor quality performance during production post-validation.

Detection of abnormal expression and splicing depends on the reference range of the target in the tested tissue, which is influenced by both the abundance and variability of the target gene expression. The commonly used metric for expression level, TPM, falls short in precisely estimating the clinical reference range. For example, compared to *TRIP11, PPP3CA* has a higher TPM in fibroblasts (31.78 versus 20.02) but a less sensitive assay reference range (0.72-1.26 versus 0.81-1.21). The wide reference range of 0.72-1.26 predicts that the expected positive finding with an expected fold change at 0.81 falls within the interval of background noise, resulting in false-negative detection. Interestingly, *PPP3CA* has a more sensitive reference range in our blood assay (0.85-1.16), enabling the detection of this challenging expression reduction in blood. A similar trend of a more sensitive reference range in blood compared to fibroblasts is observed for many other genes (**Data S6 and S7**, **Figure 3A**). In the case of the *DNM1* variant, although gene-level expression is adequate, reference range analysis reveals insufficient sequencing coverage at the junction region for the expected abnormal splicing, which explains the false-negative result for the splice junction.

The rate of NMD is a factor that can also influence diagnostic performance. When it occurs at high efficiency, NMD degrades abnormal junctions, making it easier to detect expression outliers, but harder to detect splicing outliers. This explains the FN aberrant splicing detection of the *MIPEP* junction in fibroblast and the *DNM1* junction in blood. When NMD occurs at low efficiency, expression outliers may become too modest to be detected. Our RNA-seq results revealed different fold changes for expression reduction of *AP4M1* between fibroblasts and blood, suggesting that NMD is incomplete in the blood, contributing to false-negative expression results in blood (**Figure S6**). The low expression level at the junction site in blood contributed to the false negative detection of aberrant splicing. Further understanding of tissue-specific NMD rates will guide future design and interpretation of the RNA-seq tests.

The diagnostic capability of RNA-seq tests is influenced by several technical factors, including but not limited to, sample collection methods, cell culture techniques, cDNA synthesis, library preparation, sequencing read length, sequencing depth, bioinformatics pipelines, and the configuration of control datasets (**Table 3**). Although many of these variables were not specifically examined in the current study, they warrant careful consideration and standardization during the clinical implementation of RNA-seq.

Preanalytical variables can be introduced at various stages, such as during sample collection, handling, and cell culture. The use of blood collection tubes, for example, can affect RNA integrity, yield, and gene expression profiles, potentially reducing the consistency of gene expression data across different runs ^28,29^. Similarly, variations in skin biopsy collection, such as differences in the collection site, can lead to discrepancies in gene expression ^30^. Additionally, different culturing techniques and the intrinsic characteristics of the cultured cells, such as passage number and metabolomic status, can also influence gene expression.

While it is essential for clinical labs to standardize protocols to control these variables, these challenges suggest that developing a gold standard for gene expression benchmarks based on precise read count values may not be feasible due to the high level of noise introduced by lab-specific practices in cell line strains, RNA extraction, and library preparation. To address this issue, we tentatively defined the gene expression benchmark using a binary classification of expressed or unexpressed genes. This approach allowed us to derive positive and negative gene sets from the short-read data of the reference sample GM24385 from GIAB for benchmarking. Although this method provides a lower-resolution characterization of expression levels, it enables inter-laboratory performance comparisons. As a complementary assessment, precise read count values were utilized in the reproducibility evaluation, allowing clinical labs to ensure consistent experimental and analytical procedures across replicates without relying solely on benchmark data.

Post-analytical variables can arise from sequencing experiments and data analysis procedures. Bias related to base composition, transcript strandedness, and coding versus noncoding characteristics can be introduced during cDNA synthesis and library preparation ^31,32^. Additionally, sequencing factors such as read length and sequencing depth play crucial roles in influencing test performance. Although we did not validate long-read sequencing, we utilized long-read RNA-seq data from GM24385 to construct the junction-level benchmark due to its ability to capture full-length transcripts ^33^. The extended read length facilitates accurate identification and quantification of splice junctions, enabling the detection of complex alternative splicing events that resulted in FN junctions during validation (**Data S4**, **Figure S4**).

Regarding sequencing depth, we opted for a higher overall throughput (∼150 million reads per sample) compared to previous RNA-seq diagnostic studies ^5–7^. This decision was prompted by our separate investigation, which highlighted the benefits of conducting RNA-seq at higher depth [Zhao et al., manuscript in preparation]. We found that more genes and isoforms can be detected at higher depths, and the number keeps increasing even at a depth of 1 billion reads. The rate of positive findings increases with sequencing depth, suggesting a higher clinical sensitivity at higher depth. The number of total outliers detected also increases with sequencing depth, suggesting a decreasing specificity.

In data analysis, it has been shown that decisions related to the bioinformatics pipeline, such as the choice of reference genome build version, impact the clinical performance of RNA-seq ^34^. Importantly, the robustness of outlier analysis depends on the sample size and homogeneity of the control cohort. Further research is needed to determine an optimum control sample size and to prioritize critical factors for homogeneity, such as gender and age.

In conclusion, our study provides a paradigm and necessary resources for independent laboratories to validate a clinical RNA-seq test.

## Data and code availability

Raw RNA-seq data from the validation cohort have been deposited in the NCBI Sequence Read Archive (SRA) under accession number PRJNA1124992.

## Acknowledgments

The research was supported by National Institutes of Health Common Fund (U01HG007709 and U01HG007942) and the grant from National Human Genome Research Institute (R35HG011311).

