## Supplementary material for "Clinical validation of RNA sequencing for Mendelian disorder diagnostics": Table 1

**Table 1. Sample collection and design of clinical validation**

| Sample type | | Count | |
| --- | --- | --- | --- |
| Blood | | 55 | |
|  | O&F | | 10 |
|  | Negative sample | | 24 |
|  | Positive sample | | 20 |
|  | Repoducibility | | 1 |
| Fibroblast | | 76 | |
|  | O&F | | 10 |
|  | Negative sample | | 43 |
|  | Positive sample | | 20 |
| LCL | | 2 | |
|  | Standard reference (GM24385） | | 1 |
|  | Repoducibility (K562) | | 1 |
| Total | | 130 | |

Abbreviations: O&F, optimization and familiarization

**Table 2. Analytical performance evaluation on GM24385 (HG002)**

| Repeat | Expression sensitivity | Expression specificity | Splicing sensitivity | Splicing specificity |
| --- | --- | --- | --- | --- |
| R1 | 0.9993 (0.9985-0.9997) | 1 (0.997-1) | 0.9976 (0.997-0.998) | 0.9983 (0.9966-0.9992) |
| R2 | 0.9993 (0.9985-0.9997) | 1 (0.997-1) | 0.9978 (0.9972-0.9982) | 0.999 (0.9976-0.9996) |
| R3 | 0.9993 (0.9985-0.9997) | 1 (0.997-1) | 0.9967 (0.9961-0.9972) | 0.999 (0.9976-0.9996) |
| R4 | 0.999 (0.9981-0.9995) | 1 (0.997-1) | 0.9976 (0.9971-0.9981) | 0.9979 (0.9959-0.9989) |
| R5 | 0.9991 (0.9982-0.9995) | 1 (0.997-1) | 0.9964 (0.9958-0.997) | 0.9986 (0.9969-0.9993) |

We established provisional expression and splicing benchmarks using short-read and long-read RNA-seq data of the GM24385 (HG002) lymphoblastoid sample from the Genome in a Bottle Consortium. The benchmarks are based on short-read and long-read sequencing data from independent laboratories. At the expression level, the benchmark comprises 8,991 genes positively expressed and 1,296 genes negatively expressed. At the splicing level, the benchmark includes 38,110 positive junctions and 4,195 negative junctions. Sensitivity is measured by the detection rate of positively identified genes or junctions, while specificity is measured by the proportion of negatively identified genes or junctions. Both sensitivity and specificity for expression and splicing were calculated across five experimental repeats, R1 through R5. The Wilson score method was used to calculate 95% confidence intervals for performance metrics.

Table 3. Factors that may impact the sensitivity and specificity of RNA-seq test.

| Factor | Description | Impact on result | Recommendation |
| --- | --- | --- | --- |
| Sample collection | For blood sample, the type of tube used; For fibroblast samples, the site of skin biopsy. | Different sample collection methods may introduce methodological variability, impacting statistical results. | Standardize the sample collection method. |
| Cell culture | For fibroblast samples: culture conditions, passage number, and circadian rhythm before RNA extraction. | Variations in cell conditions may result in technical variation, influencing the statistical outcomes. | Culture fibroblast cells under consistent conditions before RNA extraction. |
| cDNA synthesis method | The protocol and kit used for cDNA synthesis before sequencing. | Different cDNA synthesis methods can introduce biases (e.g., base composition bias, strand-specificity issues) that impact gene expression quantification and splicing detection. | Choose a cDNA synthesis method that minimizes bias and aligns with the specific goals of the RNA-seq experiment. |
| Library preparation | The use of polyA or ribosome depletion kits during library preparation. | Ribosome depletion kits may capture more pre-mRNA, resulting in lower sensitivity in junction analysis. | Use polyA kit for coding RNA and ribosome depletion kit for non-coding RNA. |
| Read length | Insert size and read length of RNA-seq. | Longer read lengths may improve sensitivity and specificity in junction analysis. | Opt for longer read lengths when possible. |
| Sequencing depth | Sequencing throughput of RNA-seq. | Higher sequencing depth may increase sensitivity but reduce specificity. | Balance sequencing depth with cost considerations and sequencing noise. |
| Bioinformat-ics pipeline | The software and algorithms used for aligning, normalizing, analyzing RNA-seq data, and the choice of genome build. | The bioinformatic pipeline, including genome build selection, can significantly impact gene expression estimates, detection of splicing events, and overall reproducibility and interpretation of results. | Select bioinformatic tools and pipelines based on their performance characteristics, suitability for the dataset, and robustness across different genome builds. |
| Control dataset | The control RNA-seq dataset used for outlier analysis. | Larger control sample sizes may enhance sensitivity. Heteogeneity in the reference dataset can affect outlier analysis results. Unknown or undetected shared ancestry between case and controls. | Use a larger control sample size and improve homogeneity between the test and control datasets. |
