## Supplementary material for "Clinical validation of RNA sequencing for Mendelian disorder diagnostics": Document S1

### Supplementary figures

**Figure S1. Interpretation workflow of expression and splicing outliers**

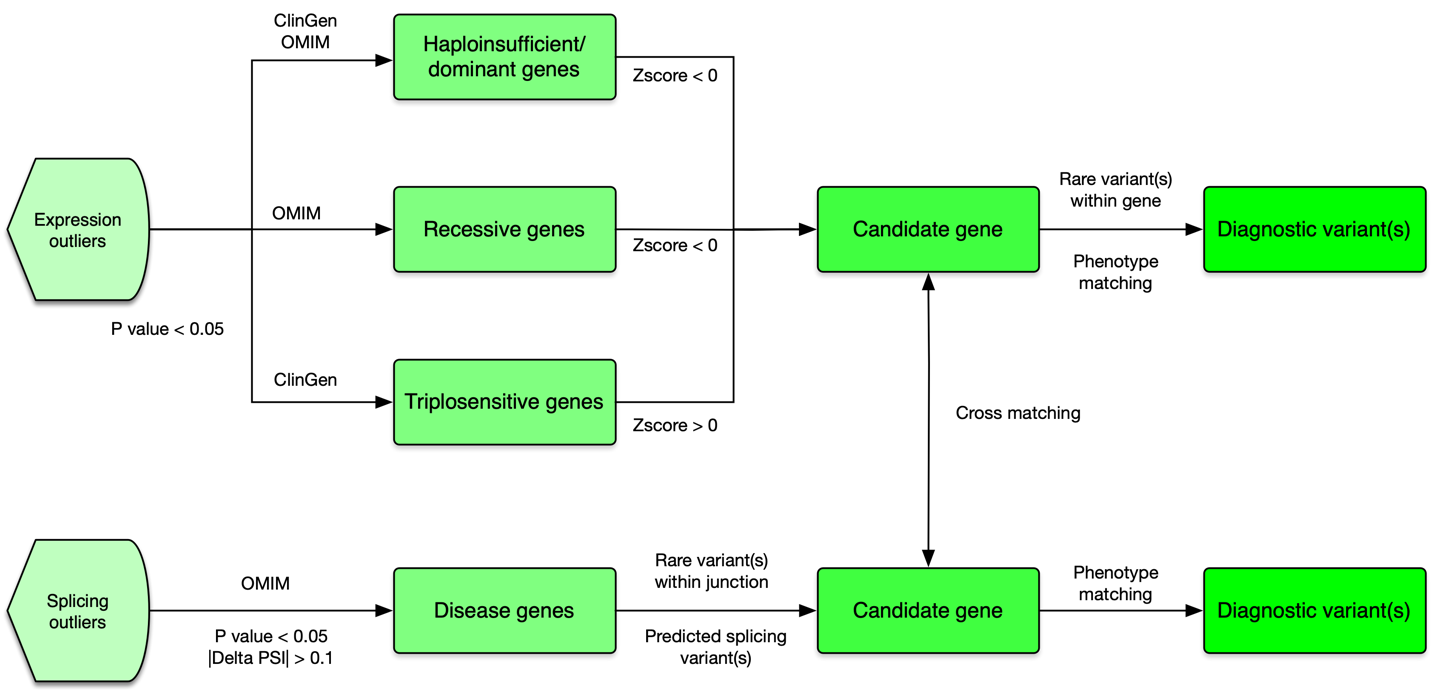

We first filter expression by P value and splicing outliers by P value and delta proportion of splice-in delta PSI. We then intersect expression and splicing outliers by OMIM and clingen disease genes. For expression outliers, we look for down-regulated genes (Zscore < 0) for haploinsufficient and recessive genes and up-regulated genes (Zscore > 0) for triplosensitive genes. We then look for rare variants within the gene that may underlie the expression outlier. For splicing outliers, we focus on splicing events with rare variant(s) located near the junction. After prioritization of candidate disease genes, we perform phenotype matching and determine whether the candidate disease gene(s) are associated with the phenotype(s) of the patient.

**Figure S2. Reproducibility analysis of K562**

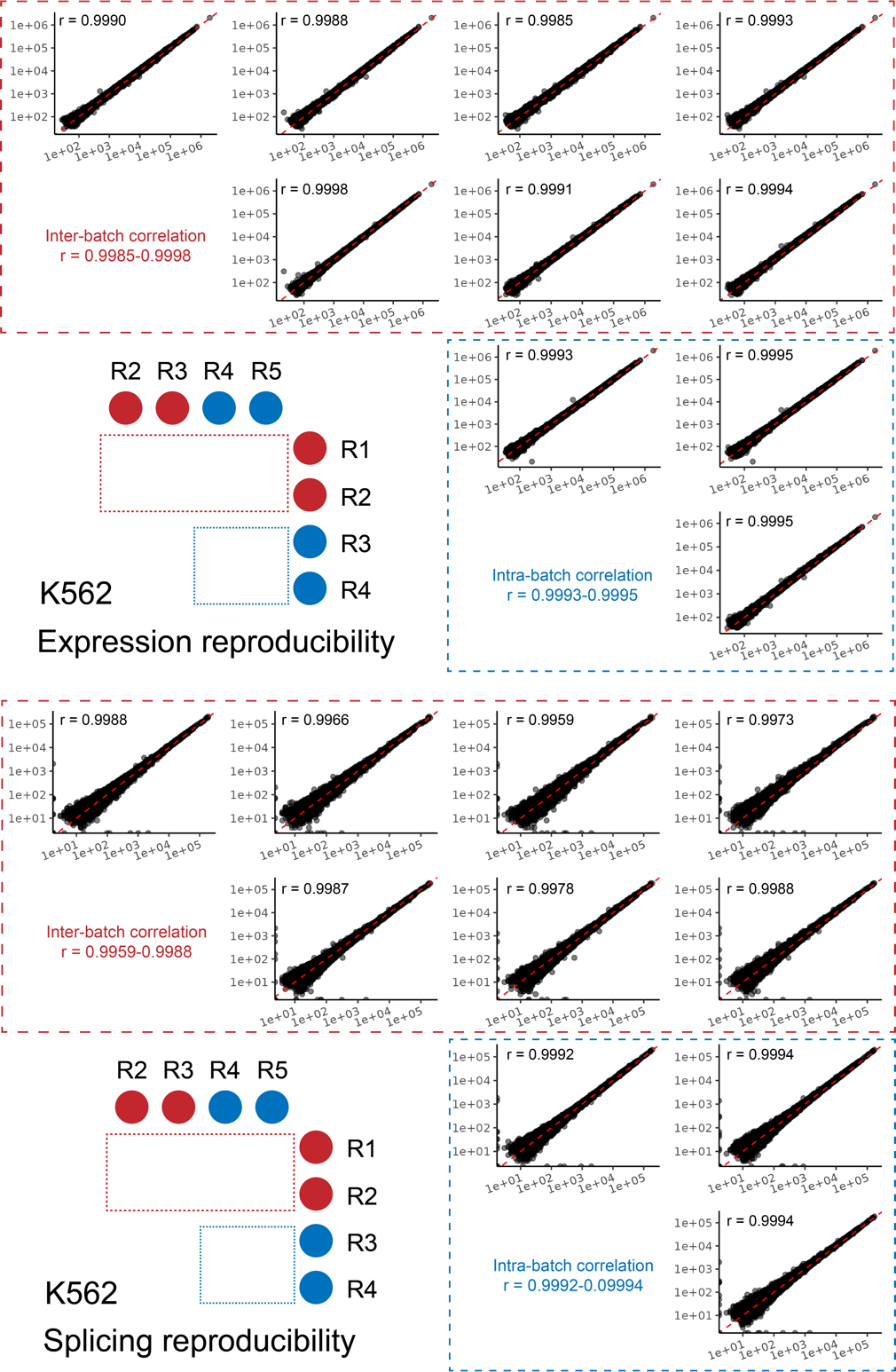

K562 was derived from the bone marrow of a patient with chronic myeloid leukemia. We conducted an intra-batch run with triplicate preparations (R3-R5) followed by two inter-batch runs (R1-R2). The expression-level correlation between two replicates was calculated using the read count of coding genes with transcript per million (TPM) > 5. The splicing-level correlation between two replicates was calculated using the read counts at GENCODE canonical junctions in genes with TPM > 5.

**Figure S3. Reproducibility analysis of BG1477**

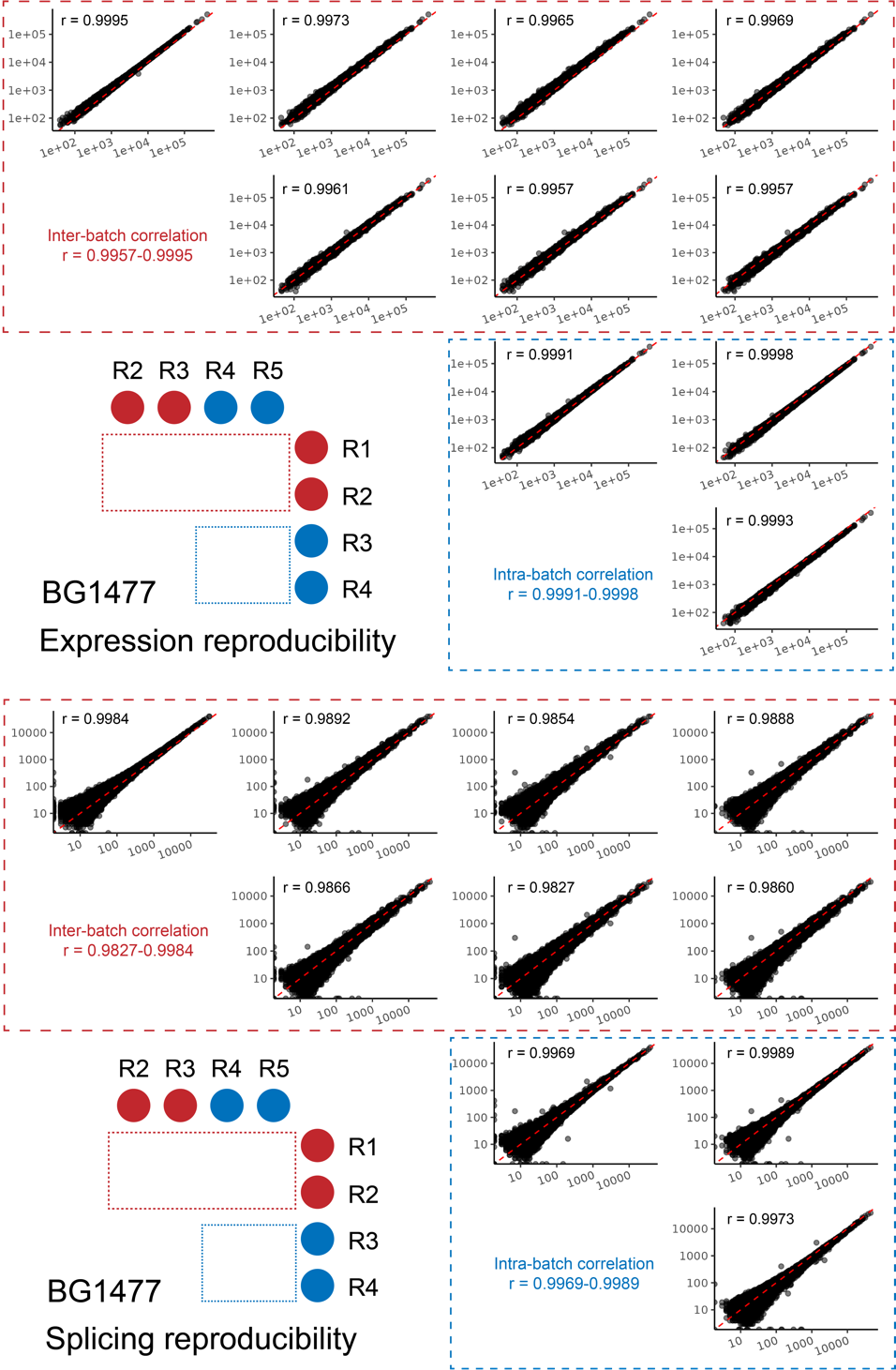

BG1477 is an in-house blood sample from a de-identified healthy donor.We conducted an intra-batch run with triplicate preparations (R3-R5) followed by two inter-batch runs (R1-R2). The expression-level correlation between two replicates was calculated using the read count of coding genes with transcript per million (TPM) > 5. The splicing-level correlation between two replicates was calculated using the read counts at GENCODE canonical junctions in genes with TPM > 5.

**Figure S4. False negative junctions in reference sample GM24385 (HG002)**

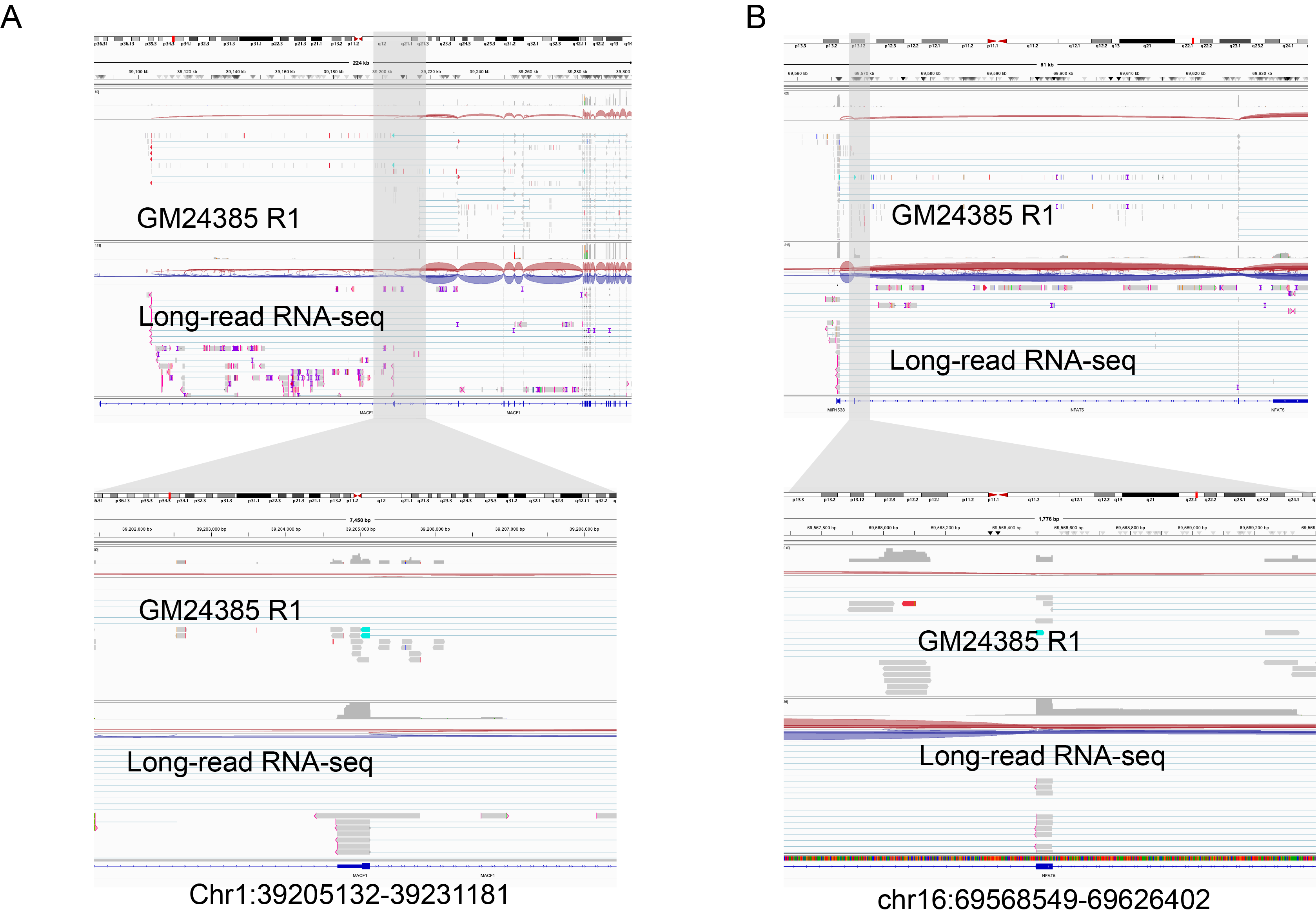

Two examples of false negative junctions in GM24385 (HG002). One is caused by an alternative start exon in *MACF1* (A). The other is caused by an alternative splicing event in *NFAT5* (B).

**Figure S5. False positive junctions in reference sample GM24385 (HG002)**

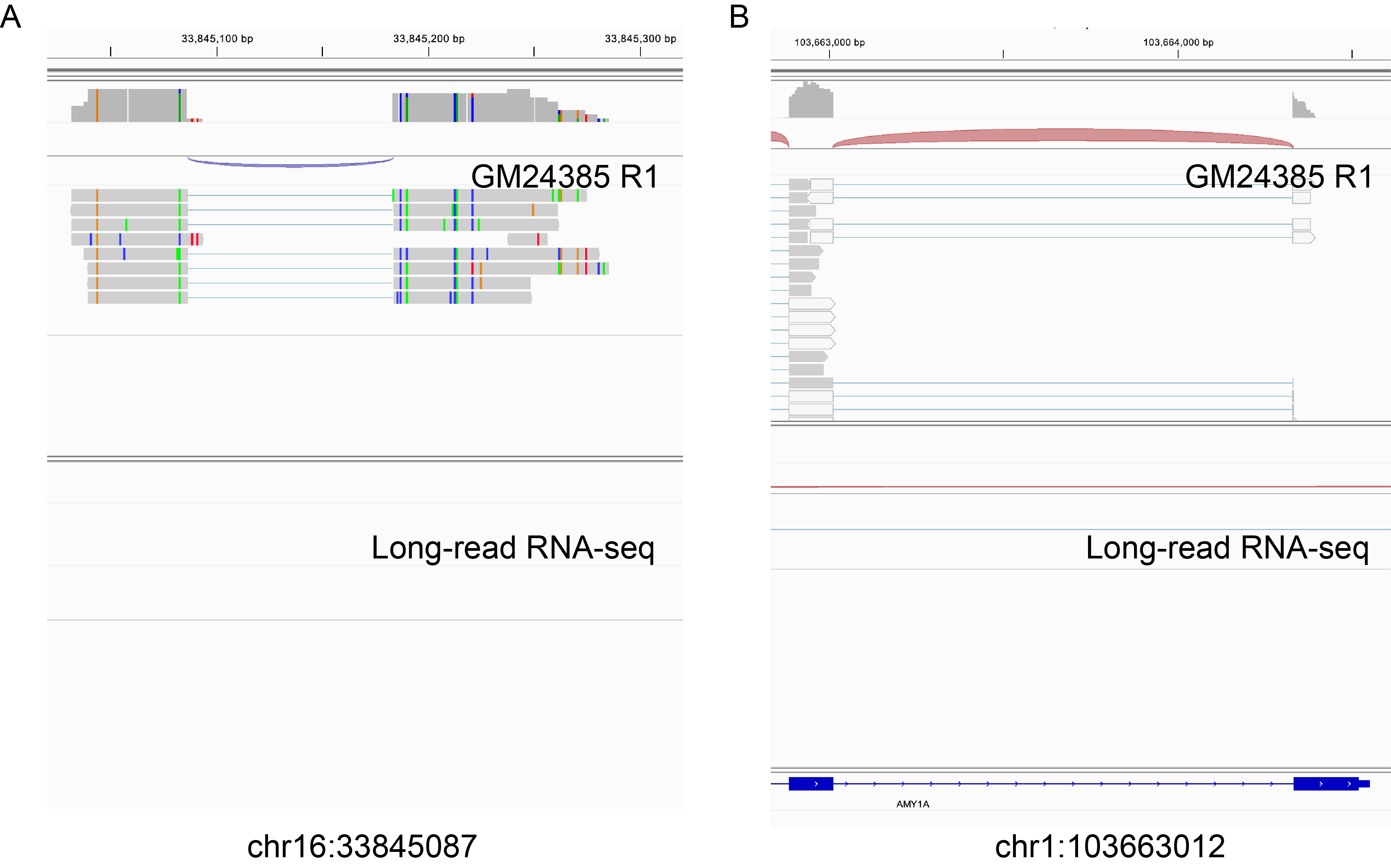

Two examples of false positive junctions in GM24385 (HG002). One is caused by mis-mapped reads in a non-coding region (A). The other is caused by mis-mapped reads in homologous genes (B).

**Figure S6. Aberrant splicing event in *AP4M1* missed in the blood sample from UD2401P0029**

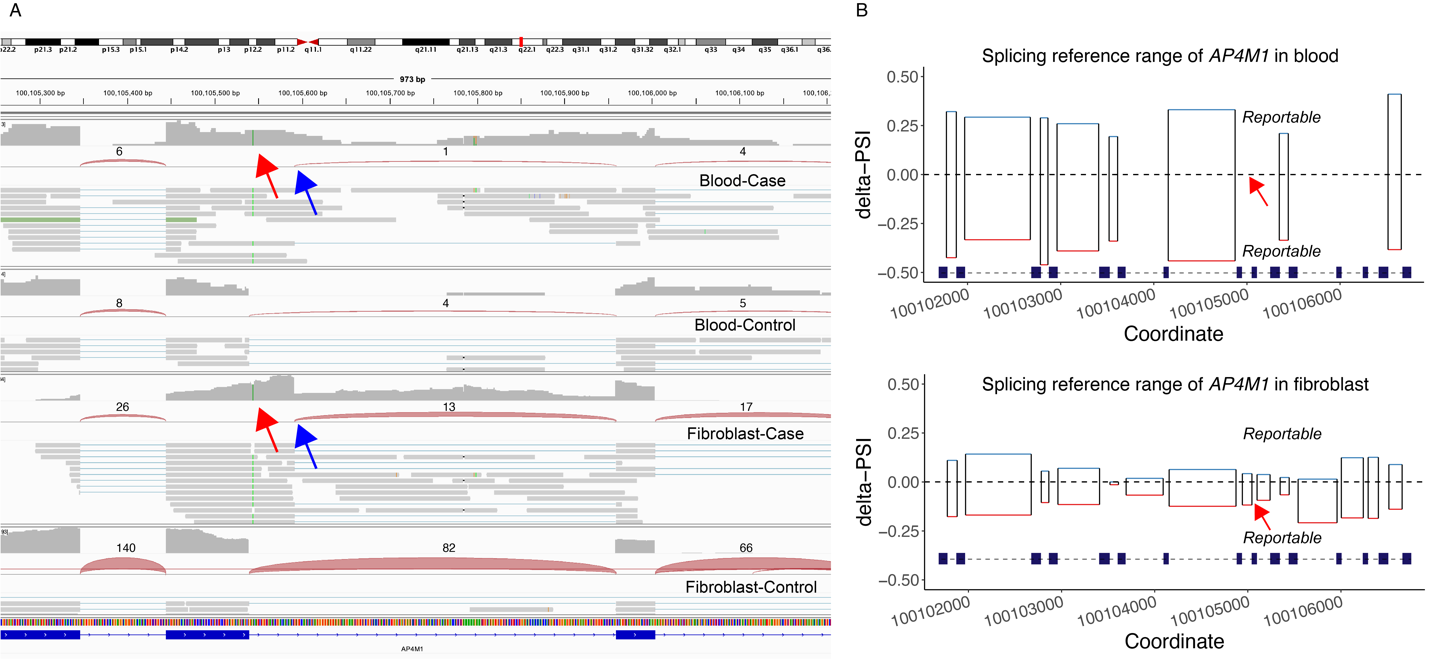

(A) A homozygous pathogenic variant NM_004722.4:c.929+5G>A in *AP4M1* (red arrows) leads to a cryptic splice donor (blue arrows), which has many supportive splitting reads in Fibroblast RNA-seq. However, only one supportive read is seen in blood RNA-seq, which is probably due to nonsense-mediated decay in addition to the low expression level of *AP4M1* in blood (TPM=2.873). (B) the splicing reference ranges of *AP4M1* based on our fibroblast and blood reference panels are shown. At the site of the pathogenic variant (red arrows), the reference range for blood was not applicable due to the low coverage, which can explain the negative result of *AP4M1* in blood.

**Figure S7. Sensitivity analysis of expression and splicing outlier identification**

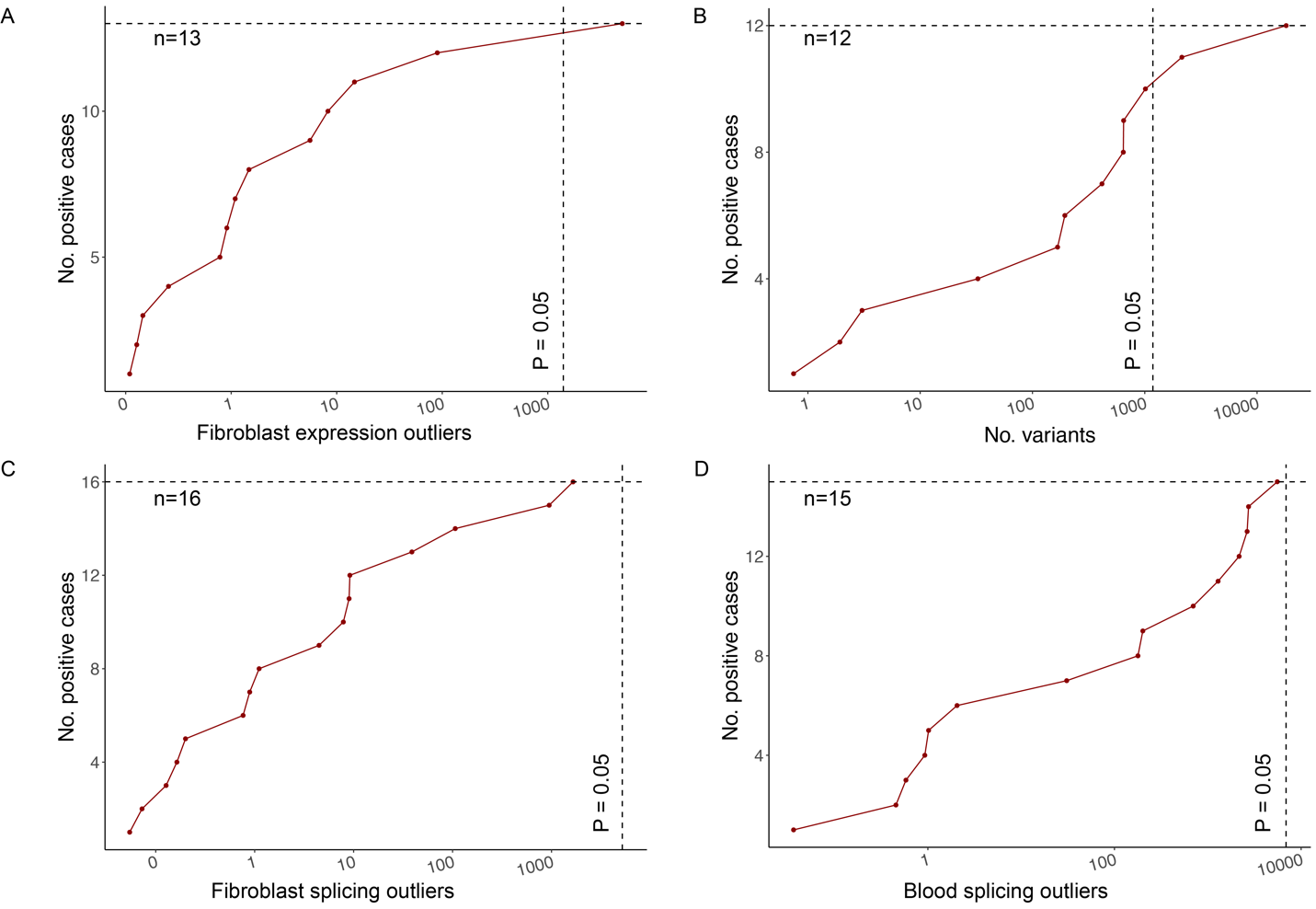

We show the relationship between the number of outliers and the number of positive cases successfully detected by the outliers at different P-value cutoffs. Expression or splicing outlier analyses in fibroblast or blood are shown in (A-D). Horizontal dashed lines indicate the total number of positive cases. Vertical dashed lines indicate the number of outliers at P-value = 0.05.
